# The association between prolonged SARS-CoV-2 symptoms and work outcomes

**DOI:** 10.1101/2024.03.10.24304054

**Authors:** Arjun Venkatesh, Huihui Yu, Caitlin Malicki, Michael Gottlieb, Joann G. Elmore, Mandy J. Hill, Ahamed H. Idris, Juan Carlos C. Montoy, Kelli N. O’Laughlin, Kristin L. Rising, Kari A. Stephens, Erica S. Spatz, Robert A. Weinstein, the INSPIRE Group

**Affiliations:** Department of Emergency Medicine, Yale School of Medicine, New Haven, Connecticut, United States of America; Center for Outcomes Research and Evaluation (CORE), Section of Cardiovascular Medicine, Yale School of Medicine, Connecticut, United State of America; Department of Emergency Medicine, Rush University Medical Center, Chicago, Illinois, United States of America; Division of General Internal Medicine and Health Services Research, David Geffen School of Medicine at the University of California, Los Angeles (UCLA), Los Angeles, California, United States of America; Department of Emergency Medicine, UTHealth Houston, Houston, Texas, United States of America; Department of Emergency Medicine, University of Texas Southwestern Medical Center, Dallas, Texas, United States of America; Department of Emergency Medicine, University of California, San Francisco, San Francisco, California, United States of America; Departments of Emergency Medicine and Global Health, University of Washington, Seattle, Washington, United States of America; Department of Emergency Medicine, Sidney Kimmel Medical College, Thomas Jefferson University, Philadelphia, Pennsylvania, United States of America; Center for Connected Care, Thomas Jefferson University, Philadelphia, Pennsylvania, United States of America; Department of Family Medicine, University of Washington, Seattle, Washington, United States of America; Department of Biomedical Informatics and Medical Education, University of Washington, Seattle, Washington, United States of America; Department of Epidemiology, Yale School of Public Health, New Haven, Connecticut, United States of America; Section of Cardiovascular Medicine, Yale School of Medicine, New Haven, Connecticut, United States of America; Division of Infectious Diseases, Department of Internal Medicine, Rush University Medical Center, Chicago, Illinois, United States of America; Department of Medicine, Cook County Hospital, Chicago, Illinois, United States of America

**Author notes:** Corresponding author (AKV). These authors are co-senior authors (Erica S. Spatz, Robert A. Weinstein). Membership of the INSPIRE Group is provided in Appendix 1.

## Abstract

While the early effects of the COVID-19 pandemic on the United States labor market are well-established, less is known about the long-term impact of SARS-CoV-2 infection and Long COVID on employment. To address this gap, we analyzed self-reported data from a prospective, national cohort study to estimate the effects of SARS-CoV-2 symptoms at three months post-infection on missed workdays and return to work. The analysis included 2,939 adults in the Innovative Support for Patients with SARS-CoV-2 Infections Registry (INSPIRE) study who tested positive for their initial SARS-CoV-2 infection at the time of enrollment, were employed before the pandemic, and completed a baseline and three-month electronic survey. At three months post-infection, 40.8% of participants reported at least one SARS-CoV-2 symptom and 9.6% of participants reported five or more SARS-CoV-2 symptoms. When asked about missed work due to their SARS-CoV-2 infection at three months, 7.1% of participants reported missing ≥10 workdays and 13.9% of participants reported not returning to work since their infection. At three months, participants with ≥5 symptoms had a higher adjusted odds ratio (aOR) of missing ≥10 workdays (2.96, 95% CI 1.81-4.83) and not returning to work (2.44, 95% CI 1.58-3.76) compared to those with no symptoms. Prolonged SARS-CoV-2 symptoms were common, affecting 4-in-10 participants at three-months post-infection, and were associated with increased odds of work loss, most pronounced among adults with ≥5 symptoms at three months. Despite the end of the Federal COVID-19 Public Health Emergency and efforts to “return to normal”, policymakers must consider the clinical and economic implications of the COVID-19 pandemic on people’s employment status and work absenteeism, particularly as data characterizing the numerous health and well-being impacts of Long COVID continue to emerge. Improved understanding of risk factors for lost work time may guide efforts to support people in returning to work.

## Introduction

The COVID-19 pandemic has resulted in tremendous economic dislocation in labor markets, creating historically volatile unemployment and reduced labor force participation rates due to unprecedented occupational health stresses and work loss [1]. Even as labor markets have stabilized in most countries including sustained periods of low unemployment [2], the economic impacts of the COVID-19 pandemic persist for many individuals as infections, hospitalizations and morbidity from SARS-CoV-2 infection continue.

Most prior research has examined the macroeconomic employment effects of the pandemic and policy responses such as stay-at-home orders [3-4], with less investigation of the direct relationship between SARS-CoV-2 illness and work. One small retrospective study conducted early in the pandemic examining work outcomes among clinical cohorts reported that approximately half of individuals hospitalized with SARS-CoV-2 were unable to return to work at six months [5]. A more recent study using the US Current Population Survey found that work absences of up to one week due to acute SARS-CoV-2 illness were associated with less labor force participation and more work absences one year later, which is estimated to have reduced the US labor force by 500,000 people [6].

One possible mechanism for this long-term impact on employment is post-COVID conditions, which include a wide range of physical and mental health consequences that are present at least four weeks after SARS-CoV-2 infection [7-8]. Post-COVID conditions, often referred to as Long COVID, affect nearly one-in-five adults with a history of SARS-CoV-2 infection [9] and may make returning to work or seeking employment more difficult [10]. An unadjusted analysis of an international convenience sample recruited via social media found that most individuals with SARS-CoV-2 symptoms beyond 28 days reported a reduced work schedule, suggesting a relationship between prolonged symptoms following an acute SARS-CoV-2 infection and short-term work loss [11]. However, the prevalence of Long COVID and its impact on work outcomes, such as return to work and missed workdays, is poorly understood [12].

To address this gap, we sought to utilize data from the Innovative Support for Patients with SARS-CoV-2 Infections Registry (INSPIRE) study to describe self-reported work outcomes related to acute SARS-CoV-2 infection and the presence of symptoms at three months post-infection.

## Materials and methods

### Study design

INSPIRE is a previously described prospective study designed to assess long-term symptoms and outcomes among persons with COVID-like illness who tested positive versus negative for SARS-CoV-2 at study enrollment [13]. Participants were enrolled virtually or in-person across eight study sites, including Rush University (Chicago, Illinois), Yale University (New Haven, Connecticut), the University of Washington (Seattle, Washington), Thomas Jefferson University (Philadelphia, Pennsylvania), the University of Texas Southwestern (Dallas, Texas), the University of Texas, Houston (Houston, Texas), the University of California, San Francisco (San Francisco, California) and the University of California, Los Angeles (Los Angeles County, California). Inclusion criteria included age ≥18 years, fluency in English or Spanish, self-reported symptoms suggestive of acute SARS-CoV-2 infection at time of testing (e.g., fever, cough), and testing for SARS-CoV-2 with an FDA-approved/authorized molecular or antigen-based assay within the preceding 42 days. Exclusion criteria included inability to provide consent, being lawfully imprisoned, inability of the study team to confirm the result of the index diagnostic test for SARS-CoV-2, having a previous SARS-CoV-2 infection >42 days before enrollment, and lacking access to an internet-connected device (e.g., smartphone, tablet, computer) for electronic survey completion. Participants with a positive SARS-CoV-2 test (COVID-positive) and a negative SARS-CoV-2 test (COVID-negative) were recruited in a 3:1 ratio. Following electronic consent, participants completed a baseline survey and follow-up surveys every three months for up to 18 months post-enrollment, although only baseline and three-month follow-up surveys were included in this secondary analysis. The three-month survey was sent 76 days following enrollment, which was up to 118 days after the positive SARS-CoV-2 test, and participants had a 28-day window to complete the three-month survey.

### Surveys

The baseline and three-month surveys included a variety of questions regarding sociodemographics, SARS-CoV-2 symptoms and overall health to assess long-term symptoms and outcomes related to COVID-like illness. To establish baseline employment status, the baseline survey asked, “Were you employed before the coronavirus outbreak?”, with the following response options: Yes; No. To establish return to work status following a SARS-CoV-2 infection, the three-month survey asked, “Did you return to work after your COVID-19 like symptoms?”, with the following response options: Yes, full-time; Yes, part-time or modified work; No; Not applicable. To establish workdays missed due to SARS-CoV-2 infection, the three-month survey asked, “Since before you had COVID-19 like symptoms, how many workdays or weeks did you miss because of health reasons?”, with the following response options: I don’t work; 0-5 workdays; 6-10 workdays; 10-20 workdays; up to 4 weeks.

To assess SARS-CoV-2 symptoms at the time of survey completion, the baseline and three-month surveys asked, “Do you currently have any of the following ongoing symptoms? (Select all that apply)”, with the following response options: fever; feeling hot or feverish; chills; repeated shaking with chills; more tired than usual; muscle aches; joint pains; runny nose; sore throat; a new cough, or worsening of a chronic chough; shortness of breath; wheezing, pain or tightness in your chest; palpitations; nausea or vomiting; headache; hair loss; abdominal pain; diarrhea (>3 loose/looser than normal stools/24 hours); decreased smell or change in smell; decreased taste or change in taste; none of the above.

### Analysis

This analysis was restricted to COVID-positive participants who responded “yes” to being employed prior to the pandemic on the baseline survey and completed the three-month survey. Unadjusted comparisons in baseline characteristics and outcomes were made using chi-square tests among groups with different numbers of symptoms (0, 1-2, 3-4, ≥5 symptoms). We examined the association of the number of symptoms at three-months with return to work and health-related work absenteeism (days missed ≥10) between enrollment and three-months using logistic regression models adjusting for age, gender, race, ethnicity, income, marital status, education, clinical comorbidity count, and SARS-CoV-2 variant time period [14]. Covariates were included in the model based on existing literature and unadjusted tests on the significance of association with outcomes. To categorize missed workdays, we established a cutoff of greater than or equal to ten days based on the original design of survey response options. All statistics and data analysis were performed in SAS 9.4.

## Results

### Survey Completion

Among 8,950 individuals who completed informed consent between December 2020 and August 2022, 6,049 were eligible for the three-month survey (**Figure 1**). A total of 4,588 participants completed the three-month survey, with survey completion rates varying slightly between the COVID-positive (77%) and COVID-negative (71%) groups. Among COVID-positive three-month survey respondents only (n=3,533), we analyzed data from the 2,939 participants (83.2%) who responded “yes” to being employed prior to the pandemic on the baseline survey.

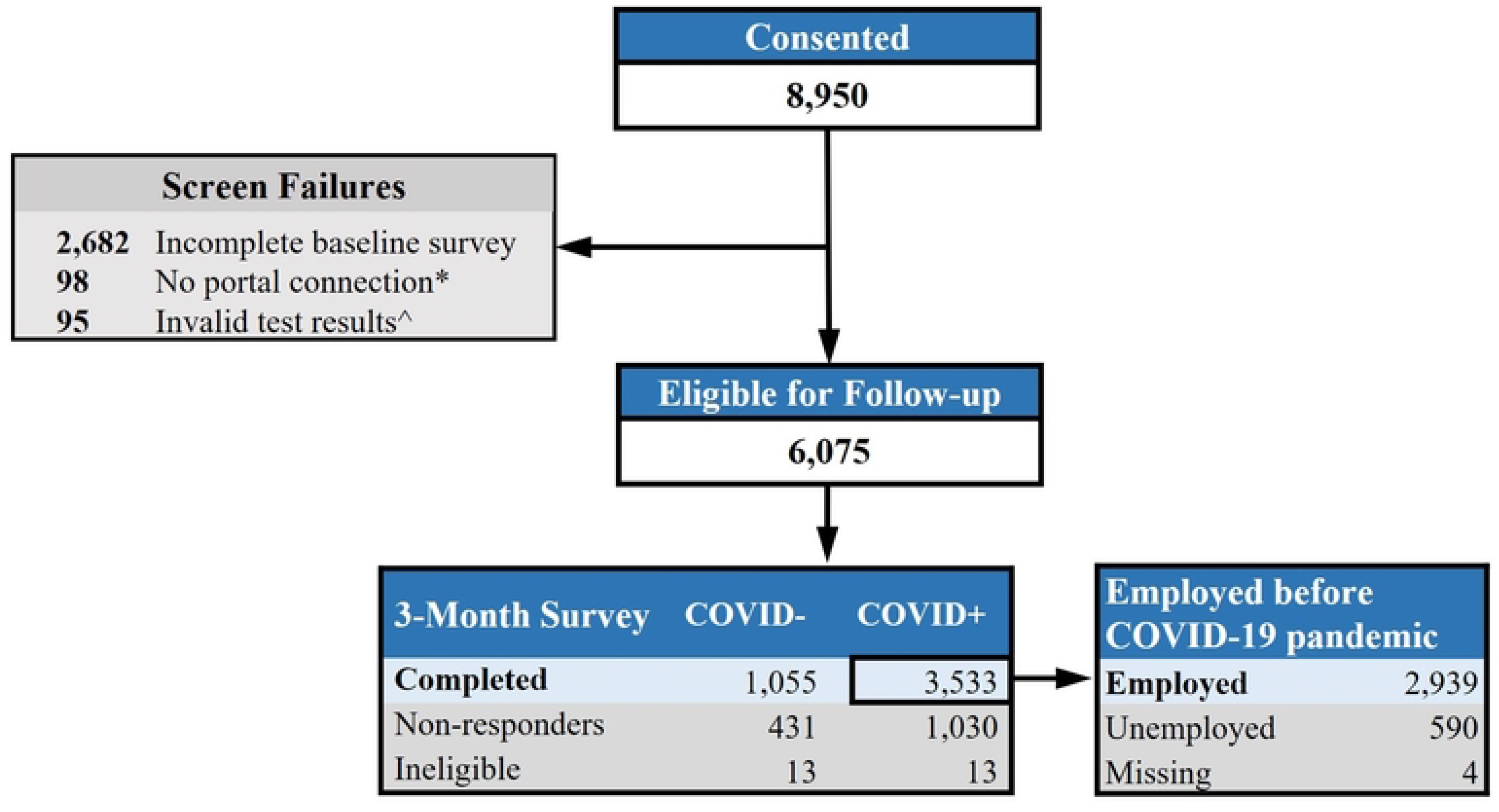

### Participant Characteristics

The mean age was 40 years (SD 12.6), 64.1% were female, 69.5% were white, 61.2% were vaccinated for SARS-CoV-2 before index test, and 3.8% were hospitalized for SARS-CoV-2 infection. 1,732 (59.2%) and 282 (9.6%) reported 0 and ≥5 symptoms at three months, respectively (**Table 1**).

| Characteristic<br>n (%) | Total<br>n=2928* | Number of Symptoms at 3 months (n, %) |  |  |  | p-value |
| --- | --- | --- | --- | --- | --- | --- |
| | | 0<br>(1732, 59.2) | 1-2<br>(677, 23.1) | 3-4<br>(237, 8.1) | $\geq 5$<br>(282, 9.6) | |
| Age |  |  |  |  |  | 0.306 |
| 18-34 | 1210 (41.3) | 750 (43.7) | 269 (40) | 87 (37) | 104 (37) |  |
| 35-49 | 1019 (34.8) | 581 (33.8) | 249 (37.1) | 88 (37.4) | 101 (35.9) |  |
| 50-64 | 548 (18.7) | 314 (18.3) | 121 (18) | 49 (20.9) | 64 (22.8) |  |
| 65+ | 129 (4.4) | 73 (4.2) | 33 (4.9) | 11 (4.7) | 12 (4.3) |  |
| Missing^ | 22 (0.8) |  |  |  |  |  |
| Gender |  |  |  |  |  | <.001 |
| Female | 1878 (64.1) | 1052 (62.5) | 458 (69.4) | 165 (72.7) | 203 (73.3) |  |
| Male | 930 (31.8) | 612 (36.4) | 191 (28.9) | 58 (25.6) | 69 (24.9) |  |
| Transgender/Non-Binary/Other | 39 (1.3) | 19 (1.1) | 11 (1.7) | 4 (1.8) | 5 (1.8) |  |
| Missing^ | 81 (2.8) |  |  |  |  |  |
| Ethnicity |  |  |  |  |  | 0.024 |
| Non-Hispanic | 2485 (84.9) | 1494 (87.9) | 565 (85) | 190 (81.5) | 236 (85.5) |  |
| Hispanic | 388 (13.3) | 205 (12.1) | 100 (15) | 43 (18.5) | 40 (14.5) |  |
| Missing^ | 55 (1.8) |  |  |  |  |  |
| Race |  |  |  |  |  | <.001 |
| American Indian/Alaskan Native | 18 (0.6) | 11 (0.7) | 2 (0.3) | 1 (0.4) | 4 (1.4) |  |
| Asian/Native Hawaiian/Pacific Islander | 389 (13.3) | 249 (14.7) | 92 (14) | 24 (10.5) | 24 (8.7) |  |
| Black | 189 (6.5) | 103 (6.1) | 32 (4.9) | 26 (11.4) | 28 (10.1) |  |
| White | 2036 (69.5) | 1217 (72) | 474 (72.3) | 148 (64.9) | 197 (71.1) |  |
| Other | 220 (7.5) | 111 (6.6) | 56 (8.5) | 29 (12.7) | 24 (8.7) |  |
| Missing^ | 76 (2.6) |  |  |  |  |  |
| Education |  |  |  |  |  | <.001 |
| Less than High school | 24 (0.8) | 11 (0.6) | 5 (0.8) | 4 (1.7) | 4 (1.4) |  |
| High school graduate | 129 (4.4) | 64 (3.8) | 28 (4.2) | 20 (8.7) | 17 (6.1) |  |
| Some College | 355 (12.1) | 185 (10.9) | 75 (11.3) | 31 (13.4) | 64 (22.9) |  |
| 2-year degree | 189 (6.5) | 88 (5.2) | 47 (7.1) | 27 (11.7) | 27 (9.6) |  |
| 4-year degree | 1018 (34.8) | 621 (36.6) | 228 (34.4) | 74 (32) | 95 (33.9) |  |
| More than 4-year degree | 1156 (39.5) | 728 (42.9) | 280 (42.2) | 75 (32.5) | 73 (26.1) |  |
| Missing^ | 57 (1.9) |  |  |  |  |  |
| Marital Status |  |  |  |  |  | <.001 |
| Married | 1665 (56.9) | 994 (57.4) | 397 (58.7) | 124 (52.8) | 150 (53.2) |  |
| Divorced/Widowed/Separated | 253 (8.6) | 123 (7.1) | 58 (8.6) | 37 (15.7) | 35 (12.4) |  |
| Never married | 1007 (34.4) | 615 (35.5) | 221 (32.7) | 74 (31.5) | 97 (34.4) |  |
| Missing^ | 3 (0.1) |  |  |  |  |  |
| Annual Family Income |  |  |  |  |  | <.001 |
| <10,000 | 77 (2.6) | 45 (2.6) | 14 (2.1) | 5 (2.1) | 13 (4.6) |  |
| 10,000-34,999 | 271 (9.3) | 137 (7.9) | 59 (8.7) | 36 (15.2) | 39 (13.8) |  |
| 35,000-49,999 | 285 (9.7) | 132 (7.6) | 79 (11.7) | 31 (13.1) | 43 (15.2) |  |
| 50,000-74,999 | 391 (13.4) | 217 (12.5) | 102 (15.1) | 32 (13.5) | 40 (14.2) |  |
| 75,000+ | 1751 (59.8) | 1105 (63.8) | 393 (58.1) | 125 (52.7) | 128 (45.4) |  |
| Prefer not to answer | 152 (5.2) | 96 (5.5) | 29 (4.3) | 8 (3.4) | 19 (6.7) |  |
| Missing^ | 1 (0) |  |  |  |  |  |
| Health Insurance |  |  |  |  |  | <.001 |
| Private | 2378 (81.2) | 1460 (84.3) | 549 (81.1) | 177 (74.7) | 192 (68.1) |  |
| Public | 383 (13.1) | 188 (10.9) | 89 (13.1) | 38 (16) | 68 (24.1) |  |
| Private and public | 68 (2.3) | 40 (2.3) | 11 (1.6) | 11 (4.6) | 6 (2.1) |  |
| None | 99 (3.4) | 44 (2.5) | 28 (4.1) | 11 (4.6) | 16 (5.7) |  |
| Housing |  |  |  |  |  | 0.450 |
| Unstable housing | 25 (0.9) | 12 (0.7) | 6 (0.9) | 4 (1.7) | 3 (1.1) |  |
| Stable housing | 2898 (99) | 1719 (99.3) | 671 (99.1) | 233 (98.3) | 275 (98.9) |  |
| Missing^ | 5 (0.1) |  |  |  |  |  |
| Tobacco Use |  |  |  |  |  | <.001 |
| Any tobacco use | 385 (13.1) | 221 (12.8) | 68 (10) | 40 (16.9) | 56 (19.8) |  |
| No tobacco use | 2541 (86.8) | 1510 (87.2) | 609 (90) | 196 (83.1) | 226 (80.1) |  |
| Missing^ | 2 (0.1) |  |  |  |  |  |
| Comorbidities |  |  |  |  |  |  |
| Asthma | 331 (11.3) | 156 (9.1) | 78 (11.6) | 44 (19) | 53 (19.6) | <.001 |
| Hypertension | 330 (11.3) | 175 (10.2) | 72 (10.7) | 34 (14.7) | 49 (18.1) | <.001 |
| Diabetes | 118 (4) | 69 (4) | 27 (4) | 13 (5.6) | 9 (3.3) | 0.613 |
| Obesity | 763 (26.1) | 397 (23.2) | 179 (26.7) | 93 (40.3) | 94 (34.8) | <.001 |
| Emphysema/COPD | 47 (1.6) | 22 (1.3) | 16 (2.4) | 3 (1.3) | 6 (2.2) | 0.219 |
| Heart conditions | 28 (1) | 16 (0.9) | 6 (0.9) | 2 (0.9) | 4 (1.5) | 0.844 |
| Kidney disease | 6 (0.2) | 2 (0.1) | 1 (0.1) | 0 (0) | 3 (1.1) | 0.008 |
| Liver disease | 15 (0.5) | 4 (0.2) | 6 (0.9) | 2 (0.9) | 3 (1.1) | 0.075 |
| Missing^ | 44 (1.5) |  |  |  |  |  |
| COVID-19 Testing Location |  |  |  |  |  | <.001 |
| At home | 469 (16) | 312 (18.1) | 117 (17.3) | 19 (8) | 21 (7.5) |  |
| Walk in clinic | 361 (12.3) | 198 (11.5) | 73 (10.8) | 34 (14.3) | 56 (20.1) |  |
| Emergency department | 73 (2.5) | 29 (1.7) | 14 (2.1) | 11 (4.6) | 19 (6.8) |  |
| Hospital | 218 (7.4) | 121 (7) | 51 (7.6) | 22 (9.3) | 24 (8.6) |  |
| Drive up testing site | 1621 (55.4) | 974 (56.4) | 375 (55.6) | 131 (55.3) | 141 (50.5) |  |
| Other | 177 (6) | 94 (5.4) | 45 (6.7) | 20 (8.4) | 18 (6.5) |  |
| Missing^ | 9 (0.4) |  |  |  |  |  |
| Hospitalization for SARS-CoV-2 |  |  |  |  |  | <.001 |
| Not hospitalized | 2779 (94.9) | 1678 (97.7) | 641 (95.7) | 219 (94.8) | 241 (89.3) |  |
| Hospitalized | 110 (3.8) | 40 (2.3) | 29 (4.3) | 12 (5.2) | 29 (10.7) |  |
| Missing^ | 39 (1.3) |  |  |  |  |  |
| Vaccination Status before SARS-CoV-2 Infection |  |  |  |  |  | <.001 |
| Unvaccinated | 535 (18.3) | 288 (21.2) | 103 (19.7) | 46 (24) | 98 (38.4) |  |
| Vaccinated | 1792 (61.2) | 1070 (78.8) | 419 (80.3) | 146 (76) | 157 (61.6) |  |
| Missing^ | 601 (20.5) |  |  |  |  |  |

### Work Outcomes

For missed workdays at three months post-infection, 197 participants (6.7%) missed ≥10 workdays, 2546 participants (87.0%) missed <10 workdays, 184 participants (6.3%) were not applicable (i.e. did not have work), and 1 participant (0.0%) was missing a response. For return to work at three months post-infection, 386 participants (13.2%) did not return to work, 2388 participants (81.6%) returned to work, and 154 participants (5.3%) were missing responses. The unadjusted bivariate analyses showed that participants with ≥5 symptoms had significantly higher odds of missing ≥10 workdays (18.8%) than participants with fewer (7.6-12.7%) or no symptoms (4.5%) at three months. Similarly, the unadjusted bivariate analyses showed that participants with ≥5 symptoms were significantly more likely to not return to work (27.8%) compared to participants with fewer (15.1-16.3%) or no symptoms (10.5%) at three months.

After adjusting for participants’ sociodemographic and history of clinical conditions, participants with ≥5 symptoms had the greatest odds of missing ≥10 workdays and not returning to work compared to those with fewer or no symptoms at three months (**Figure 2**). For participants with ≥10 missed workdays, there was a “dose-response” relation to number of symptoms: Compared to participants with no symptoms at three months, the odds of missing ≥10 workdays was two times higher in participants with 3-4 symptoms (adjusted odds ratio [aOR]=1.94, 95% CI: 1.08-3.49) and nearly three times higher in participants with ≥5 symptoms (aOR=2.96, 95% CI: 1.81-4.83). The difference in odds of missed workdays was not significant among participants with 1-2 symptoms compared to those with no symptoms at three months (aOR=1.58, 95% CI: 0.998-2.49). Compared to participants with no symptoms at three months, the odds of not returning to work were higher in participants with ≥5 symptoms (aOR=2.44, 95% CI: 1.58-3.76) and in participants with 1-2 symptoms (aOR=1.74, 95% CI: 1.23-2.47). The difference in odds of not returning to work was not significant among participants with 3-4 symptoms compared to those with no symptoms at three months (aOR=1.25, 95% CI: 0.73-2.14).

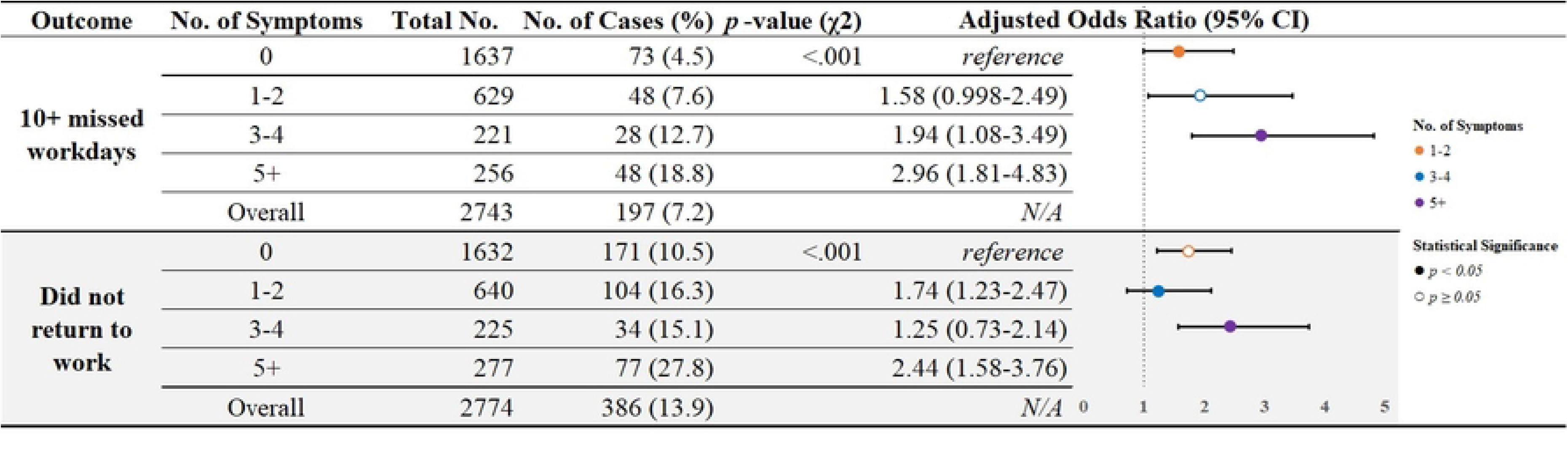

## Discussion

The presence of SARS-CoV-2 symptoms three months after acute infection was pervasive and was associated with greater odds of work loss at three months in a prospective registry of people with an initial SARS-CoV-2 infection. This association was especially pronounced among adults with a greater symptom burden (≥5 symptoms) at three months, who experienced two to three-fold increased risk of substantial missed workdays and not returning to work compared to adults whose symptoms resolved by three months.

Our work extends prior literature in a few notable ways. First, by capturing clinical information beyond the acute SARS-CoV-2 infection period and including primarily adults with milder disease, the relationships identified in the INSPIRE registry are likely more generalizable to working U.S. adults than prior work in more limited populations. In addition, because the INSPIRE registry includes baseline and longer-term follow up data, this analysis supports temporally connecting persistent SARS-CoV-2 symptoms to long-term work outcomes. Second, the consistent relationship between increased three-month symptom burden and worse work outcomes further bolsters the likelihood that health effects of post-COVID conditions are sufficient to explain the well-documented relationship between health and work status. Lastly, the broad enrollment period of the INSPIRE registry between December 2020 and August 2022 demonstrates the durability of these findings despite the likely heterogeneous effects of SARS-CoV-2 infection upon work alongside evolving variants, severity of disease, vaccination and treatment.

Given high SARS-CoV-2 vaccination rates (77% among participants with non-missing data) and the mild disease course observed in this cohort, the magnitude of work loss is striking. Extrapolating to 208 million adults working in the U.S., of whom at least 42% are estimated to have been infected with SARS-CoV-2 as of May 2022 [15], our data suggest that symptomatic SARS-CoV-2 infection may have contributed to over 12.9 million individuals not returning to work within three months of infection, of whom 2.4 million may have post-covid conditions. Given the disproportionate burden of SARS-CoV-2 infection observed among workers in public-facing industries, such as education and healthcare, the economic impacts of Long COVID may be disproportionately distributed across the nation [16].

As with all studies, this analysis has limitations. First, estimates may overattribute work loss due to SARS-CoV-2 as our analysis did not include COVID-negative participants and other pandemic-related causes. Second, our analyses considered each symptom equally, which may not capture nuanced relationships between symptom type, symptom severity and work outcomes. Third, survey questions and responses were developed iteratively in response to the changing landscape of the pandemic, and analysis was limited by survey design and available data. Lastly, there is risk of residual confounding as we were not able to adjust for all covariates, such as disability, which were found correlated with the covariates included and outcomes.

## Conclusion

Findings suggest that prolonged SARS-CoV-2 symptoms are common, affecting 4-in 10 participants at three months post-infection and are associated with increased odds of work loss, with the most pronounced work loss association among adults with ≥5 at three months. As data characterizing the numerous health and well-being impacts of Long COVID emerge, despite efforts to “return to normal,” policymakers must consider the clinical and economic implications of the COVID-19 pandemic on people’s employment status and work absenteeism and strategies for reducing absenteeism.

## Data Availability

The existing analytic dataset includes protected health information and personally identifiable information, including dates, which cannot be shared due to privacy and confidentiality of research health information.

## Acknowledgements

We would like to thank the California Department of Public Health, CTSI COVID Clinical Research Steering Committee, CTSI Office of Clinical Research Patient Navigation Team, and Bioinformatics Program Public Health Seattle King County for their assistance with participant recruitment. We would also like to thank the University of Washington Institute of Translational Health Sciences (ITHS) for support of the REDCap instance and for biomedical informatics resources used by the UW Clinical Core and Enrolling Site to enable study recruitment, which is funded by the National Center for Advancing Translational Sciences of the National Institutes of Health under award number UL1TR002319.

## Supporting Information

**S1 Fig. This is the S1 Fig Title**. This is the S1 Fig legend.

**S2 Fig. This is the S2 Fig Title**. This is the S2 Fig legend.

**S1 Table. This is the S1 Table Title**. This is the S1 Table legend.

**S1 File. This is the S1 File Title**. This is the S1 File author group.

